# Temporal considerations in the 2021 COVID-19 lockdown of Ho Chi Minh City

**DOI:** 10.1101/2021.08.04.21261332

**Authors:** Emmanuel L. C. VI M. Plan, Huong Le Thi, Duy Manh Le, Haidang Phan

**Affiliations:** Institute of Theoretical and Applied Research, Duy Tan University, Hanoi 100 000, Vietnam; Faculty of Natural Sciences, Duy Tan University, Danang, 550 000, Vietnam; Department of Mathematics and Informatics, Thang Long University, Hanoi, Vietnam; Institute of Theoretical and Applied Research, Duy Tan University; Faculty of Pharmacy, Duy Tan University, Danang, 550 000, Vietnam; Faculty of Civil Engineering, Duy Tan University, Danang, 550 000, Vietnam

## Abstract

The success of Vietnam in controlling the spread of COVID-19 hinges on a timely implementation of its coherent strategy of containment and rapid tracing and testing efforts. The Vietnamese living in Mekong Delta are currently being besieged by the SARS-Cov-2 Delta variant as they undergo several and extended levels of lockdown. In this work we examine the temporal aspects of the lockdown in Ho Chi Minh City and predict the progress of the outbreak in terms of the total number of confirmed cases.

A compartmental model with containment is fit to data to estimate the rate of transmission in Ho Chi Minh City. The severity of the lockdown is estimated from publicly-available data on mobility and coupled to the rate of infection. Various scenarios on when to begin a lockdown and its duration are assessed. This report, dated 27 July 2021, supports a lockdown of at least 3 weeks and predicts that there could be half as many cases had the inevitable lockdown started a week earlier.

## I. INTRODUCTION

Vietnam was globally known in 2020 for its success in keeping the number of cases the 2019 Coronavirus Disease (COVID-19) arising from community transmission low [1]. Despite its geographical proximity to China and when COVID-19 was recording more than 80 million cases in 2020, the total number of cases in Vietnam, a country of more than 90 million inhabitants, was less than 1500. One of the reasons Vietnam attained this feat is that they managed to keep the outbreaks localized by timely and effective non-pharmaceutical interventions [2]. Even before the virus spread worldwide in the early 2020, the Vietnamese government already enforced lockdowns, locally known as ‘social distancing’: schools and workplaces were closed, and border entry was strictly limited (henceforth, lockdown, ‘social distancing’, or containment will be used interchangeably to refer to the same idea). These efforts kept the total number of cases in the first wave at 268 and even gave Vietnam 99 consecutive days without community transmission from April to June 2020 [3]. Similar interventions, then including rapid and mass testing utilizing locally-developed test kits [4], kept the number of confirmed cases to 962 in the second wave concentrated in Da Nang last July 2020 [3, 5]. Swift and effective localized lockdowns remained the most effective response of the Vietnamese government in the third wave, which began last January 27, 2021. In that outbreak, clusters of infection were detected in the cities of Hai Duong and Quang Ninh [6]. Within 2.5 months, Vietnam was again cleared of the virus after tallying a total of 1586 cases [7].

Lockdowns constitute of only one aspect of the approach of the Vietnamese government, which is to coordinate “a proactive containment strategy” that utilizes tracing, quarantining, and comprehensive testing [2, 4]. While the community is contained, massive testing is used to identify the network of clusters and prevent wider transmission. Core to this rapid identification of infected individuals is an easily identifiable classification system that describes an individual’s connection to a confirmed case, along with corresponding testing and quarantine measures [8]. Once a COVID patient is identified and classified as “F0”, people who might have been in contact with the patient in the past 14 days would be classified as F1 individuals and tested immediately. They would then either be in isolation for 14 days if the result is negative, or will be quarantined at a hospital if positive. Simultaneously, close contacts of F1 individuals (F2s) were required to self-isolate at home for 14 days and are required to inform authorities for testing. This system of contact tracing and testing quickly identifies areas that will have to be isolated from the community.

The 4th wave of COVID-19 outbreak in Vietnam is different from the previous three in that, as of writing, approximately 10^5^ cases have already been confirmed [9, 10], and challenging its overall strategy containment and contact tracing. In the current outbreak, the highly urbanized Ho Chi Minh City (HCMC) with approximately 9 million inhabitants is most affected. As of writing (July 27, 2021), there are approximately 72000 cases in HCMC alone. The outbreak has spread to a number of provinces in Southern Vietnam, e.g. Binh Duong (8909 cases), Long An (3931 cases), Dong Nai (2714), Dong Thap (2397 cases), Tien Giang (1825 cases), Tay Ninh (938 cases) [10]. The rapid increase in the number of new cases is attributed to the appearance of the Delta variant of the SARS-CoV-2 virus [11]. This variant is estimated to be 60% more transmissible than previous variants and even partially resistant to the vaccines in existence [12, 13].

In order to grasp the severity of this larger outbreak, the Ministry of Information and Communications in HCMC tapped the support of and provided relevant data to two research groups, one from Fulbright University (HCMC), and the other is Tech4Covid, affiliated to Vietnam National University - Ho Chi Minh University of Science [14]. The groups, using data until June 27, 2021, initially estimated that the pandemic in HCMC should have approached its peak by early July with an approximate total of 11000 cases. However, despite the city-wide semi-lockdown (Directive 15) in place as of May 31 and a full lockdown (Directive 16) in the high-risk areas (see Appendix A for details on these containment measures), the number of cases continued to increase, with at least 1000 *daily* cases starting July 9, 2021. A day after the nationwide graduation exams were held, the Vietnamese government placed the entire HCMC under Directive 16: strictly moderating inflow and outflow of residents and goods, as well as limiting movement to the most essential needs.

In this report we model the spread of COVID-19 in Ho Chi Minh City but using the compartmental models and giving emphasis on the strategy of containment. A version of compartmental models that is most well-known are the Susceptible-Exposed-Infectious-Removed/Recovered (SEIR models), in which each individual of the whole population under consideration is, at any given time, classified into these homogeneous compartments according to their health status. These models and their modifications have been widely used in studying many types of infectious diseases, including COVID-19 [15–22]. In order to quantify the isolation and tracing strategies in HCMC, we extend the classical SEIR model by adding two key compartments: quarantined (Q) and hospitalized (H). Models employing such compartments have been previously used to model the desired interventions with success [15–19]. The infection rate will be estimated from the total number of cases in HCMC, which corresponds to compartment H in the model.

To focus on the effect of a timely and effective lockdown strategy, we scale the infection rate with a time-dependent parameter *p* that is estimated from the observed mobility of the residents. While smaller values of *p* clearly translate to a smaller infection rate, it is of importance to policy makers when and for how long to impose a lockdown, especially because doing so effects huge societal and economic impact [23, 24], aside from its effects on mental health [25]. We show here that there is wisdom in the recent extension of the implementation of Directive 16 in HCMC [26] from 15 days to 3 weeks, and that the implementation of the lockdown last July 9, while could be argued to be late, also averts HCMC from a more undesirable situation.

In the next section, we detail the model and the assumptions involved. Our results and predictions are given in section III, and conclusions and recommendations are provided at the end.

## II. MODEL AND FITTING

### A. The model

Here a modified SEIR model with quarantine and hospitalized (confirmed) compartments is used to model and predict the total number of COVID-19 infected individuals in HCMC (see schematic in Fig. 1). Individuals are classified based on their status: a person is classified as susceptible (*S*) if she can be infected; exposed (*E*) if she has been infected but has yet to be infectious; infectious if she can transmit the virus to others with a further sub-classification on whether that person is asymptomatic (*I*_*a*_) or symptomatic (*I*_*s*_). Note that it is assumed that a fraction *α* = 0.2 of the infected population do not display any symptoms [27], and may recover (*R*) without being confirmed, unless they have been traced. The symptomatic individuals will definitely be hospitalized (*H*), which will be what will be compared to the total number of cases in HCMC. Lastly, exposed individuals may be quarantined (*Q*); this may be significant if contact tracing is efficient.

**FIG. 1.**
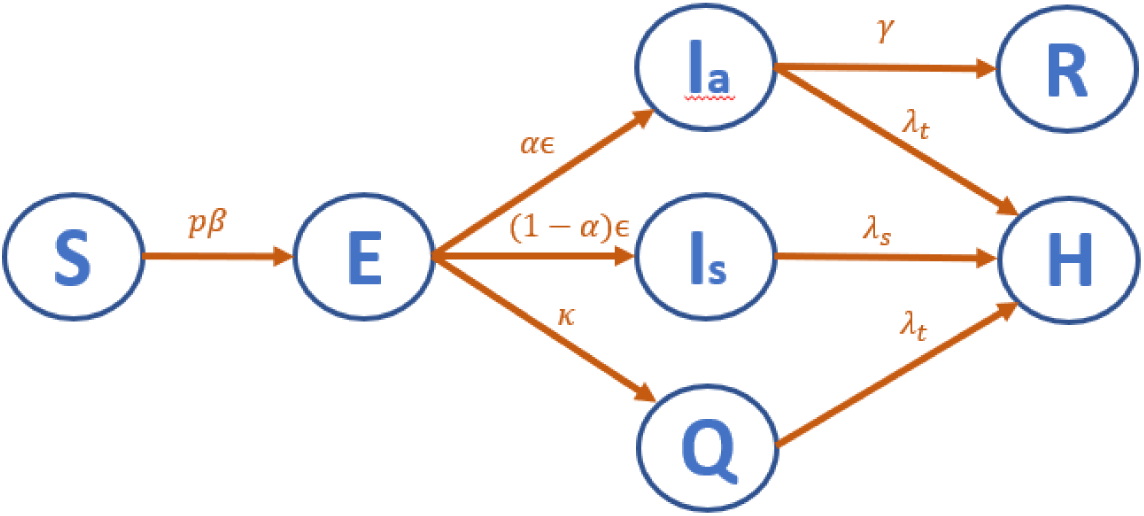
A schematic of the model used

The equations that govern the dynamics of disease spread are as follows:

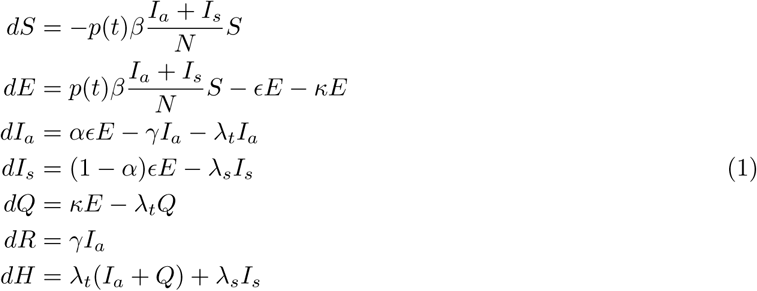

where *N* is the total population. The transition rates between compartments, as seen in Fig. 1 and the equations above, are summarized in the table below along with their values. Note that the parameter *p* = *p*(*t*) aims to capture social distancing by scaling down the infection rate [28]. Here, the value of *p* is estimated via the Google Mobility Data for Ho Chi Minh City (see Appendix B for more details). This parameter varies depending on the ‘social distancing’ restrictions imposed by the Vietnamese government, primarily through the implementation of Directives 15 and 16. The effect of an institutional intervention usually results into an epidemik peak some 9-25 days later (e.g. [29–31],); here, we assume a delay of 11 days before the value of *p* plays a role in the dynamics [14].

Symptomatic individuals are isolated and confirmed on average 1 day after developing symptoms (*λ*_*s*_ = 1, while asymptomatic and exposed individuals are isolated and confirmed at a mean rate of 2 days after being asymptomatic or exposed (*κ* = *λ*_*t*_ = 1*/*2), a day after the contacts of symptomatic individuals are traced; the latter reflects the rapid tracing efforts of Vietnam. Epidemiological characteristics of COVID-19 allowed the authors to estimate the incubation rate as *E* = 1*/*4 and *γ* = 1*/*10. The incubation rate of the Delta variant, while yet to be fully quantified, is estimated to be faster than the original variant, and the recovery rate of mild cases is known to be less than 10 days [32, 33]. Hence, only the parameter *β* is estimated from the cumulative case numbers via the compartment H.

For this model we ignore natural birth and death rates. It must be noted that this model assumes that isolated and confirmed individuals are perfectly quarantined and cannot infect others any longer. Immunity through vaccination is also not accounted for in this model, in spite of the recent efforts of the Vietnamese government to inoculate as many individuals in HCMC as possible [34]. The timeframe of this study (70 days) will not allow a significant percentage of the population to be immune due to limited supply of vaccines and the natural delay of developing antibodies. Preliminary results of an agent-based vaccination model of an urban city in Vietnam show that vaccination as a response to an outbreak will have negligible effect during the first month, will not significantly affect the duration of the outbreak, but may decrease the number of total infections after 3 months if a sufficient fraction of the population is inoculated [35]. Lastly, this model isolates HCMC and does not count infections in nearby provinces which may have originated from HCMC. Given that the infection has spread in the neighboring provinces, this work underestimates the real force of infection in the southern region of Vietnam.

### B. Fitting and Evolution

The infection rate *β* is first estimated by fitting Eqs. (2), through compartment *H*, to the total number of confirmed cases in Ho Chi Minh City [36] using 15 days of data (June 23-July 7). Note that the parameter *p* is assumed to be constant during the time period of the fitting (HCMC is under semi-lockdown). Numerical integration was performed using the Dormand-Prince pair Runge-Kutta (4,5) in Matlab [37].The prediction beyond July 7 displays reasonably good fit two weeks after data used after the data fit (see blue thick curve in Fig. 2, left), with only larger deviations on July 25-26. The confidence interval of the estimated *β* is obtained and provides a range of the estimated total number of cases (pink dashed curves in Fig. 2). Note that in this figure, the lockdown is assumed to extend indefinitely in this figure, and there would have been less than 500 cases by mid-August, and no more cases by the end of that month. It must be remarked that the daily number of cases on the third week of July start to deviate from the predictions, which may either because of *p* fails to quantify effectively the lockdown efforts or that there are factors beyond mobility that affects the transmission.

**FIG. 2.**
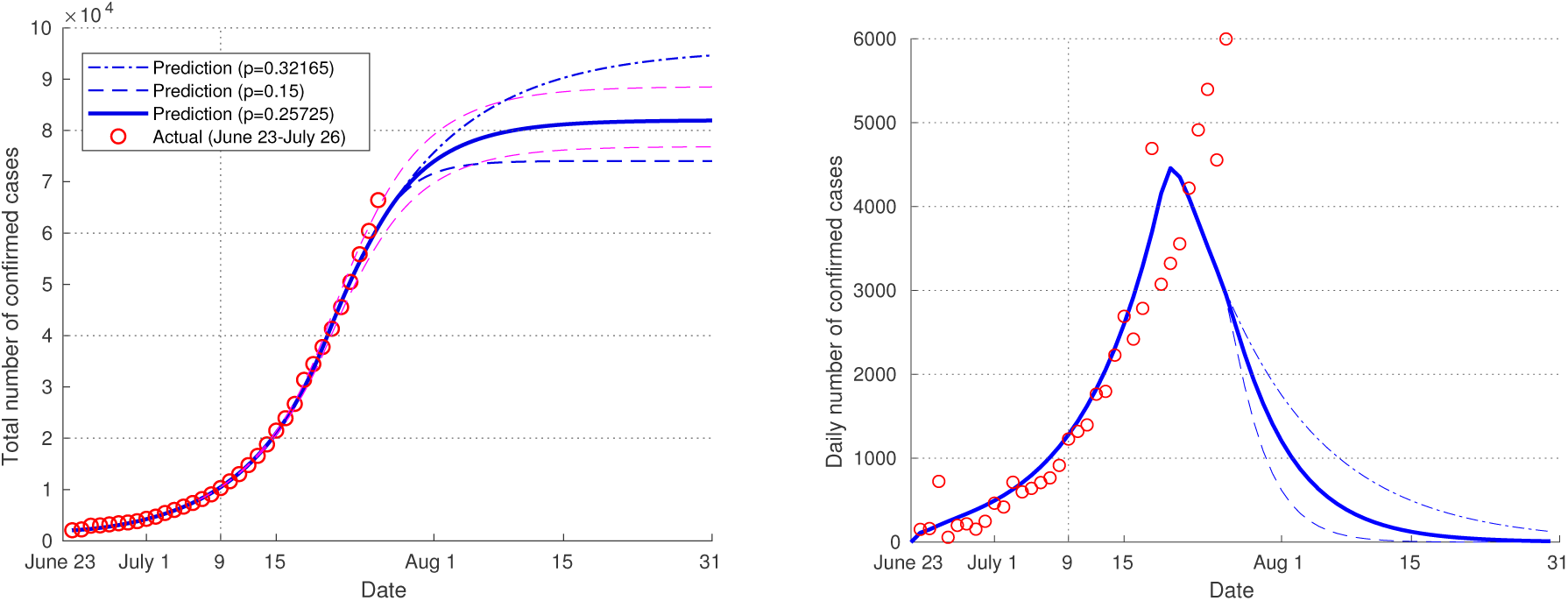
(Left) Model fit (black), data (red circles) and predictions for the total number of COVID-19 cases in Ho Chi Minh City, with Directive 16 resulting in: *p* = 0.25725 (thick straight blue curve), *p* = 0.35 (dash-dotted blue curve, theoretical), *p* = 0.15 (dashed, theoretical). In dashed pink lines are predictions using the limits of the confidence intervals for *β*. The vertical dotted line indicate the implementation of Directive 16 (July 9). (Right) Corresponding daily number of cases for the scenarios on the left.

## III. RESULTS AND DISCUSSION

### A. Effect of *p* on the total number of cases

In order to aid policy-makers and motivate science-based decisions on lockdowns, we predict the total number of cases in HCMC by evolving the equations of the compartmental model from June 23 until Aug 31 (70 days). The effect of changing *p* = *p*(*t*) as a function of the ‘social distancing’ affects the curves by segmenting the epidemic curve into several curves, most noticeable in the curve for daily infections (Fig. 2, right). By reducing the transmission rate, lower values of *p* will lead to lower number of cases. Once an intervention takes an effect, it can completely change the course of an epidemic from exponential growth to exponential decay.

To examine the critical value of *p* that determines exponential decay, we first discuss the epidemiological parameter *R*_0_, which quantifies the average number of people who get infected from a single infected person. If *R*_0_ *>* 1, the epidemic is expected to grow, while if *R*_0_ *<* 1, the situation will be controlled in time. The reproductive number *R*_0_ of the fitted model, calculated via the next generation matrix, is given as [38]:

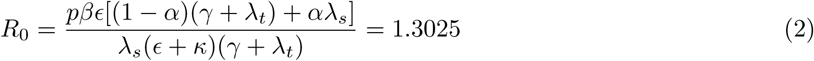

The estimated *R*_0_ = 1.3025 implies that every 100 infected persons might have spread it to around 130 other individuals. This estimate is slightly higher than the *R*_0_ = 1.22 reported by the Tech4Covid group, which used a different model and data earlier in this outbreak [39]. Note that this estimate for *R*_0_ already assumes that Directive 15 has taken an effect by lowering *p*, but this proves to be insufficient to reduce *R*_0_ below unity.

The effect of varying the intensity of the lockdown via *p* in this model is straightforward because *R*_0_ ∝ *p*. Hence, so long as mobility is limited to *p* = 0.537385*/*1.3025 ≈ 0.41258, then the pandemic will be controlled in time. The outcome when various values of *p* are enforced are shown in Fig. 2. A stricter lockdown (*p* = 0.15 *<* 0.25725) than one enforced by July 9 may not change much in terms of the cumulative number of cases, but it does allow a more controllable number earlier. In contrast, a weakly-enforced lockdown - like that during the transition from Directive 15 to 16 (*p* = 0.35 ≈ 0.32165), while still effective in the long-term, may result to at least 13000 more cases and possibly a delayed lifting of governmental interventions.

### B. Temporal aspects of implementing a lockdown

In order the understand the effect of a timely and sufficiently long lockdown, we consider scenarios where Directive 16 would have been implemented earlier/later and for different time duration. Previous research on timely lockdowns in the United States showed that a delay beyond a week before tallying 5 cases will lead a large increase in the number of cases for the next 50 days [29]. It would nevertheless be useful to assess the effectiveness of timely intervention in a developing country.

To do this, we calculate the expected total number of COVID-19 cases expected in HCMC by August 31 under different starting dates of the full lockdown and for different durations *d*. In as far as the model is concerned, lifting directive 16 must however not result into an *R*_0_ *>* 1. It is thus not sufficient to simply transition from Directive 16 to Directive 15 since *p* = 0.537385 ⇒ *R*_0_ = 1.3025 *>* 1. For our purposes, we propose a Directive that is more relaxed than Directive 16 but stricter than Directive 15 and take *p* = (0.537385 + 0.25725)*/*2 ≈ 0.397318, which give a reproductive number *R*_0_ = 0.963 *<* 1, which is a slow decrease in the daily number of cases.

Figure 3 shows a heatmap showing the relative number of total number of cases in HCMC as a function of the temporal aspects of a lockdown. For instance, had the full lockdown started on July 3 and lasted for 3 weeks before going into a partial lockdown, there would have been half as many cases than the case when the lockdown started on July 9. In contrast, delaying the start of the lockdown by 4 days would have seen a 60% increase in the total number of cases. This chart shows that there is a larger gradient along the axis representing the lockdown date, implying that early lockdown interventions result in larger decreases in the total number of cases than by extending lockdowns. By contrast, while the duration of the lockdown plays a role, it only affects by providing at least a 5% reduction for an early lockdown, and approximately 20% difference between full lockdowns of length 2 weeks and that of 5 weeks for a delayed lockdown. Indeed there is no huge benefit to extend a lockdown given a small outbreak. Nevertheless, it must be noted that around 2/3 of these decreases in the total number are achieved by lengthening a full lockdown from 15 to 21 days. For the actual situation where Directive 16 is imposed on July 9, extending the lockdown from 15 to 21 days may provide a 9% decrease, but extending it for another week may only decrease it further by 3%.

**FIG. 3.**
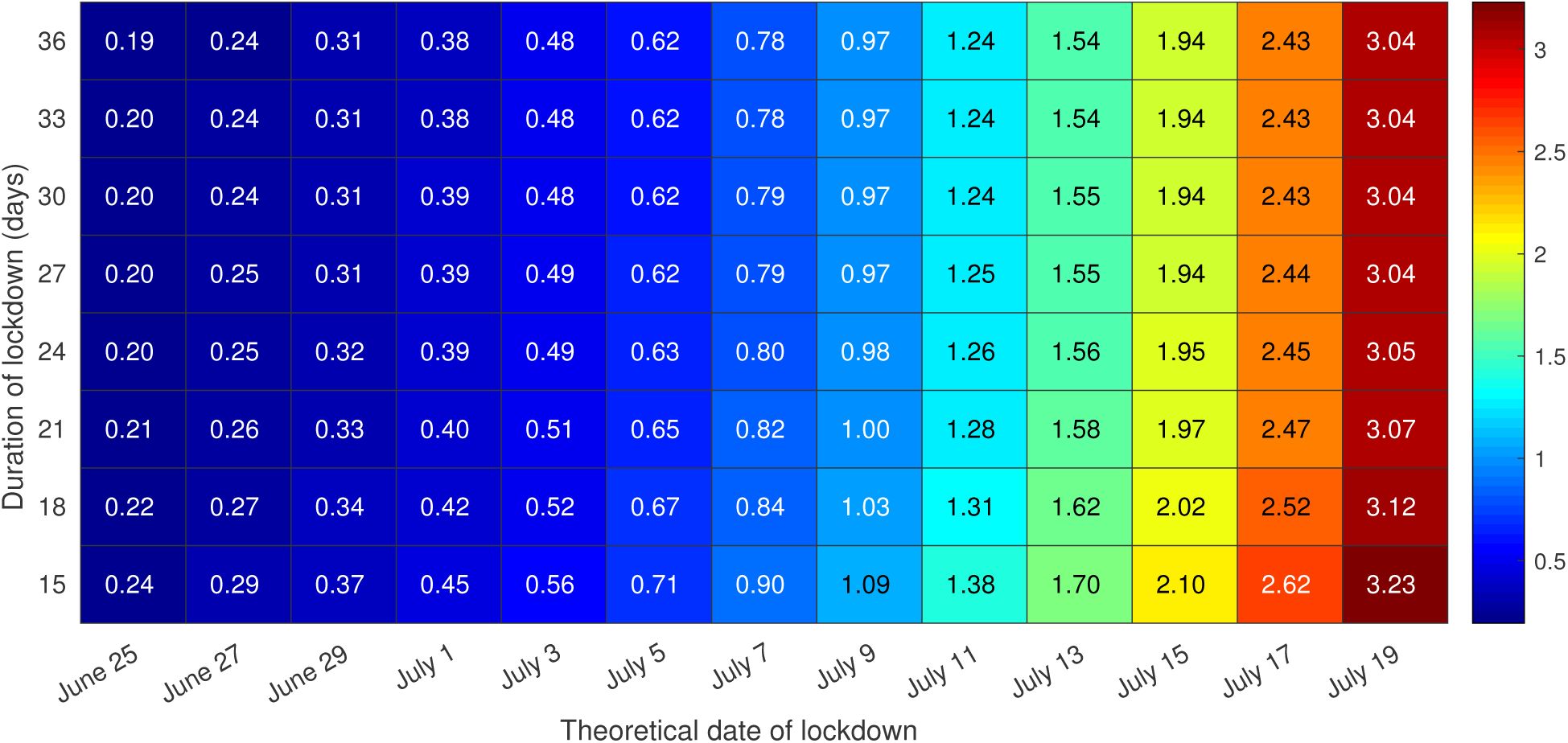
Total number of cases in HCMC by August 31 as a function of when a full lockdown would have been enforced and the duration *d* of the full lockdown, rescaled by the total number of cases given a lockdown of length *d* = 21 days starting July 9 (approximately 8.2 *×* 10^4^ cases).

Implementing a full lockdown has serious economic impacts, notwithstanding social and mental health impacts [23–25], and the Vietnamese government then has to balance the economic and societal repercussions with its necessity. Figure 4 examines more closely the 8th column of the heatmap, i.e. the effect of various duration *d* on the total/daily number of confirmed cases given the an implementation of lockdown on July 9, with the solid curve representing *d* = 21 days. Since Directive 16 will result in *R*_0_ well below 1 and assuming that its effects will be felt until 11 days after lifting, extending the full lockdown beyond 21 days (July 29) provides incremental decrease in the number of total cases by Aug 31. On the other hand, a shorter duration *d* = 15 can easily add 10000 more cases, owing to the fact that there will again be increased mobility while the number of cases are still huge (around 900/day in early August) and hence, more opportunities for the virus to spread.

**FIG. 4.**
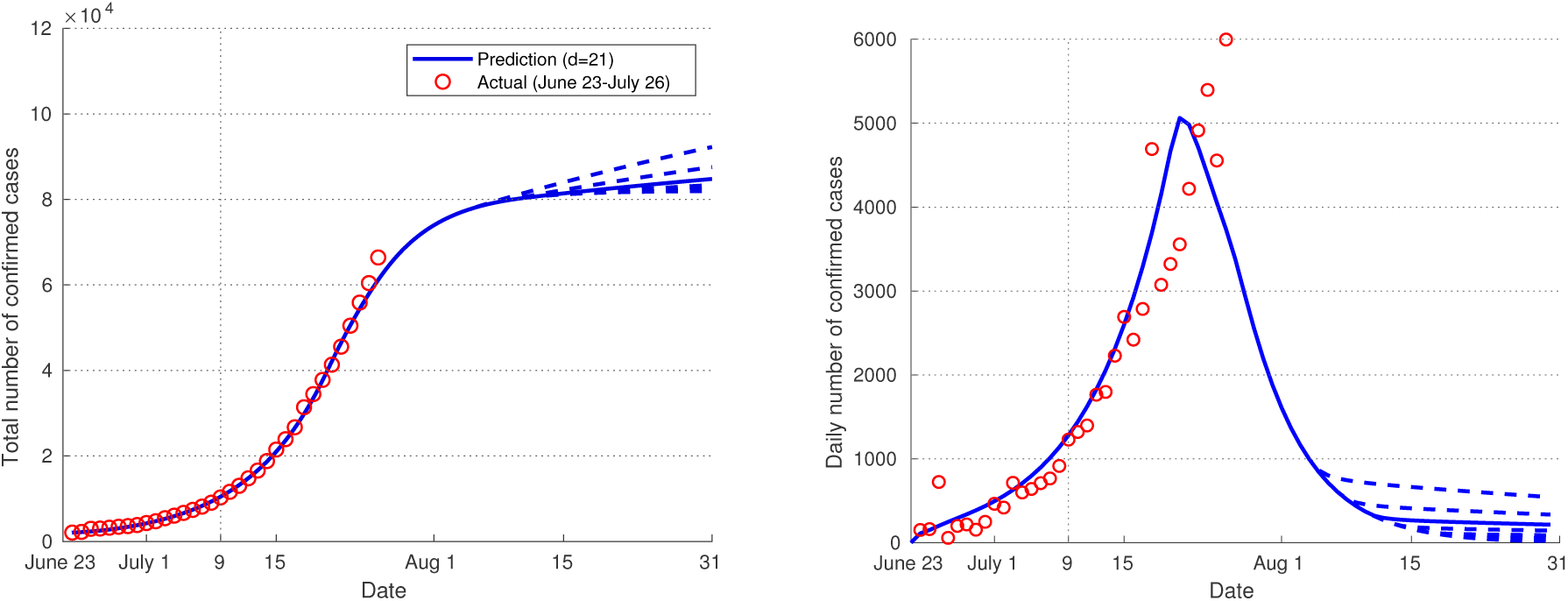
The total (left) and daily (right) number of confirmed cases as a function of the length of the lockdown *d*, from 15 to 36 days in increments of 3. The solid line denotes the case *d* = 21. Higher(lower) values of *d* give lower(higher) numbers of confirmed cases.

More than the duration of a lockdown, the heatmap above confirms that the timeliness of an intervention plays a significant role in the spread of COVID-19 [29]. In light of future outbreaks in Vietnam, we further examine the epidemic curves for hypothetical start dates of the lockdown. Figure 5 shows the total/daily number of confirmed cases for different dates of implementing Directive 16, assuming a lockdown of 21 days. Every two days of earlier lockdown could have resulted in approximately 1.5 × 10^4^ less cases (by August 31), whereas every two days beyond July 9 would have potentially resulted into at least 30000 cases more, with a delayed lockdown (July 17) reaching 200000 cases by the end of August in HCMC alone. Clearly an early and strong intervention can result into less cases and possibly a shorter ‘social distancing’, if the number of daily infections is used as an indication when to lift a lockdown. Figure 5 (right) shows that earlier lockdowns manage to reduce the number of cases to any threshold daily number earlier than later lockdowns.

**FIG. 5.**
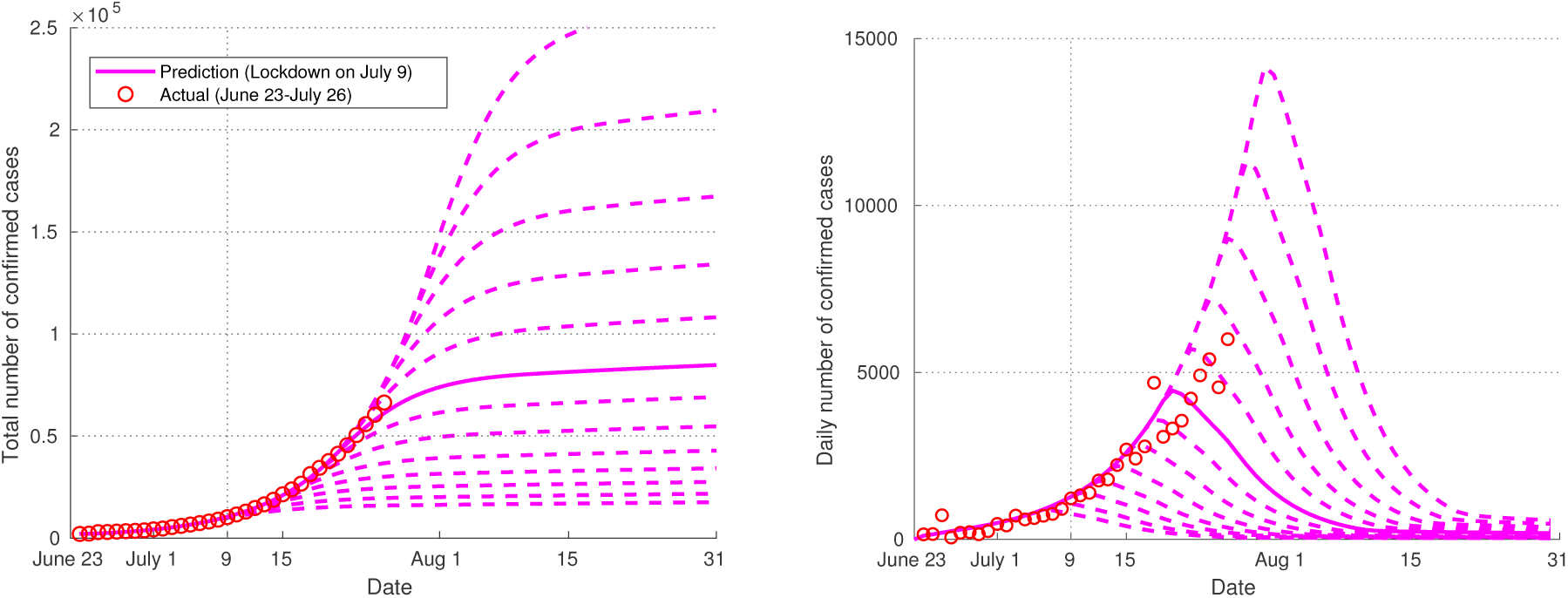
The total (left) and daily (right) number of confirmed cases as a function of the length of the lockdown of when the lockdown is enforced, in increments of 2 days (June 25 to July 19). The solid line denotes the case when it is enforced on July 9. Earlier(later) implementation of Directive 16 results in lower(higher) numbers of confirmed cases.

## IV. CONCLUSION

In this work, we model the ongoing COVID-19 outbreak in Ho Chi Minh City as a function of the containment measures of the government by controlling the transmission rate as a function of the mobility of people. The parameter *p* was used to estimate the fraction of the population mingling with others and which is proportional to the rate by which the disease spreads. A compartmental model with containment is fit with the cumulative case incidence in HCMC and the total number of cases in the coming weeks is predicted. This report also explores quantitatively the effects of the temporal aspects of implementing a lockdown on the total number of infected cases. With the goal of minimizing this number, an earlier lockdown is always advised as this avoids the exponential increase in the number of cases. Moreover, a lockdown duration of at least 3 weeks is ideal as there are noticeable improvements compared to a 15-day lockdown.

From the initial writing of this manuscript to its submission, there had been more significant deviations in the prediction of the case incidence as the assumption on the interventions already manifest, whereas the actual case incidence has yet to manifest an indisputable decline. One aspect that must be explored in more detail is the natural delay from the day of implementation of an intervention to the time that its effects manifests. Previous research shows that the delay from a lockdown to the peak in daily case incidence is widely spread from 9 days to 25 days [31], and could thus be an impediment for modellers and policy makers as they would need to assess if their efforts have sufficient impact on the further prevention of the outbreak. Here this delay was assumed to be 11 days as proposed by another Vietnamese study [14], but this may perhaps be validated in a future work, e.g. by comparing to previous outbreak, or those from other countries if to account for the same strain of virus. Another possible direction is to extract a non-trivial relationship between the mobility data to the parameter *p*, e.g. a linear approximation that varies daily.

Other aspects of studies involving the effects of lockdown would involve interdisciplinary studies in economics or in healthcare management. For instance, assuming certain economic costs in implementing (semi-or full) lockdowns as well as healthcare costs, how would this affect the timing and duration of lockdowns if the goal is to minimize macroeconomic or microeconomic costs? On the aspect of healthcare, it would be interesting to know the the timing and intensity of an intervention in order to ensure continuous quality provision of healthcare services. This study on the timely imposition and sufficient duration of lockdowns in Vietnam would certainly provide a first step for these studies.

## Data Availability

All data used are publicly available

### Appendix A: Vietnam Government Directives

The differences between Directive 15 and Directive 16 are summarized as follows [40]. While not explicitly said in the directives, ride-hailing and delivery/shipping services are suspended under Directive 16 but are allowed in Directive 15 [41]. Moreover, some offices and institutions that are allowed to operate at limited (e.g. 50%) capacity under Directive 15 are also closed under Directive 16 [42].

### Appendix B: Estimating *p* from Google Mobility Data

The data provided by Google show the change in the number of visitors in a given location (workplaces, retail and recreation, transit stations, parks, and groceries and pharmacies) or change in the duration (residential), relative to a baseline [43]. The data obtained is shown in Fig. 6. From May 31, upon the implementation of Directive 15, mobility is on average, at *v* ≈ −48.8205. From the implementation of Directive 16 on July 9, there is to be a short transitional period of 6 days during which mobility is *v* ≈ −69.3667, after which Directive 16 is in full force with mobility at *v* ≈ −75.5.

**FIG. 6.**
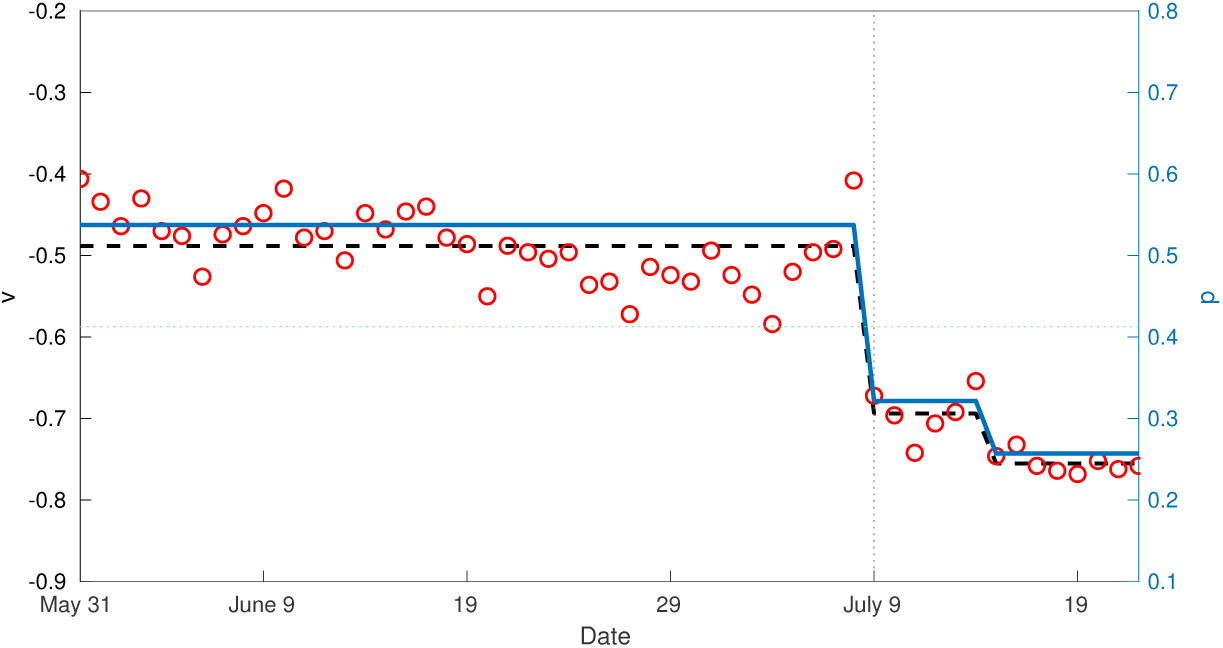
(Left scale) Average of the non-residential mobility data *v* (red dots) and the average constant value (black dashed) over the time periods described in Table I. (Right scale) The value of *p* derived from the mobility data for each time period (blue line). The green line shows the threshold value for *p* = 0.41258 that gives *R*0 = 1.

Since the baseline is based on data from January 3 to February 6 2020, during which Vietnam was celebrating Tet holiday and was implementing ‘social distancing’ measures, it is necessary to first rescale *v* in order to estimate *p*, which measures the reduction of the transmission rate. To estimate for *p*_*i*_ for day *i*, we compute

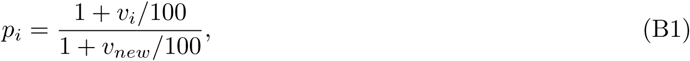

where the google mobility percentage *v*_*i*_ is the average of the non-residential mobility data. The daily values are first converted to actual values by multiplying 1 + *v*_*i*_*/*100 with a baseline value *b*, and then compared to a new baseline *b*(1 + *v*_*new*_*/*100). Here, *v*_*new*_ is taken as a three-week average from 2021 January 4 to 2021 January 24, between the New Year holiday and the Lunar New Year in February (and the third outbreak in Northern Vietnam), when mobility, particularly work, is deemed normal. The values of *p* shown in Table I were obtained by getting the averages of *p*_*i*_ over the three time periods concerned.

**TABLE I.** The parameters and their values

| Parameter | Value | Remarks |
| --- | --- | --- |
| $p$ | 0.537385 | (May 31-July 8*) Proportion of population susceptible; Estimated |
|  | 0.32165 | (July 9-July 15*) |
|  | 0.25725 | (July 16 onwards*) *not including delay |
| $\beta$ | 6.5133 | Transmission coefficient; Fitted |
| $\epsilon$ | 0.25 | Incubation rate; Assumed |
| $\kappa$ | 0.5 | Isolation rate; Assumed |
| $\alpha$ | 0.2 | Proportion of asymptomatic; Assumed |
| $\gamma$ | 0.1 | Recovery rate; Assumed |
| $\lambda_t$ | 0.5 | Tracing and confirming rate; Assumed |
| $\lambda_s$ | 1.0 | Confirming rate; Assumed |

**TABLE II.** The differences between Directive 15 and 16

| Safety measures | Directive 15 | Directive 16 |
| --- | --- | --- |
| Schools | Closed | Closed |
| Safe distance observed in public | 2m | 2m |
| Non-essential businesses | Closed | Closed |
| Essential businesses | Allowed | Allowed |
| Gathering in public (outside schools, hospitals and offices) | < 10 people | $\leq 2$ people |
| Events | $\leq 20$ people | Not allowed |
| Transport from/to areas with outbreaks | Restricted<br>(need negative COVID-test) | Not allowed |
| Public transport | Limited | Suspended |

